# Postural control telerehabilitation with a low-cost virtual reality protocol for children with cerebral palsy: Protocol for a clinical trial

**DOI:** 10.1101/2022.04.25.22274289

**Authors:** Valeska Gatica-Rojas, Ricardo Cartes-Velásquez, Alex Soto-Poblete, Luis Eduardo Cofré Lizama

**Affiliations:** Tele-rehabilitation technology center and Neurosciences in Human Movement, Faculty of Health Sciences, Universidad de Talca. Av. Lircay S/N, Talca, Chile. Email address; Fundación Kimntrum, Concepción, Chile. Email address; Institute of Mathematics and Physics, Universidad de Talca. Av. Lircay S/N, Talca, Chile. Email address; School of Allied Health, Human Services and Sports, La Trobe University, Victoria, Australia. Email address

**Keywords:** telerehabilitation, cerebral palsy, balance, virtual reality, covid-19 pandemic

## Abstract

**Objective:** To establish the feasibility and effectiveness of a rehabilitation programme using low-cost virtual reality aimed at improving postural control in children with cerebral type palsy spastic hemiplegia. It also seeks to compare low-cost virtual reality under two delivery modalities, telerehabilitation (TR) and face-to-face (FtF).

**Methods:** Randomized controlled clinical trial from the ACTRN platform ACTRN12621000117819. Eighteen sessions of low-cost virtual reality therapy will be provided through both, FtF and TR modalities using a Nintendo Wii balance board. Each programme will last for 6 weeks and will consist of 3 sessions per week of 25 minutes each. The participants will include 40 patients diagnosed with cerebral palsy type spastic hemiplegia. Twenty participants for each group. Participants will be assessed at baseline, by the end of weeks 2, 4, and 6, and at weeks 8 and 10 (post-intervention follow-ups). Clinical measures include the Modified-Modified Ashworth Scale for lower limbs, Modified Ashworth Scale for upper limbs, timed up-and-go tests, the timed one-leg standing and 6-minute walk test. Posturographic measures, including sway area and velocity, under six conditions will be used: 2 statics and 4 dynamic conditions, which include voluntary sway in the mediolateral direction following a metronome set at 30Hz and 60Hz, and sway while playing 2 different videogames.

**Results:** This study provides an assessment of the feasibility and effectiveness of an affordable rehabilitation programme using low-cost virtual reality aimed at improving postural control in children with cerebral palsy.

**Conclusion:** Rehabilitation programme using low-cost virtual reality will improve postural control in children with cerebral palsy type spastic hemiplegia and this programme delivered using TR will be as effective as a FtF modality. The TR programme has be designed to expand the coverage of physiotherapy services for children with cerebral palsy in low-resource settings and in remote areas.

## Introduction

Cerebral palsy (CP) is the most common condition treated by therapists in infant neurological units, and it is the most common cause of motor disability in children and adolescents,^1,2^ with an increasing prevalence in developing countries.^3^ According to the topographical classification, spastic hemiplegia (SHE) is one of the most prevalent types of CP.^1,4^ In SHE, the mobility of one side of the body is more affected, generating poor postural control during standing, leading to impaired performance in of daily functional activities.^5,6^ To date, most studies have focused in developing interventions to improve upper-limb function in SHE, for example, virtual reality interventions, which further allow undertaking therapeutic training at home.^7-9^ The development of therapeutic interventions aimed at improving postural control, on the other hand, has received less attention.^10-12^

Efficient, inexpensive, accessible, and engaging are some of the highlighted characteristics sought after in the development of novel physiotherapy interventions, particularly in children.^13-16^ Since the recent COVID-19 pandemic has interrupted non-essential rehabilitation services, including those focusing on patients with CP, an urgent need for remotely deliverable and less supervised interventions has also been highlighted to provide health access to underserved populations.^17^ In this regard, telerehabilitation (TR) with the use low-cost technological tools and exergames provide with an opportunity to meet the before mentioned needs and which can further expand qualitatively and quantitatively neurorehabilitation practice. This TR approach is not excluding of at-clinic interventions, which combined may allow for a flexible dual-care model that integrates hospital and home-based rehabilitation.^13^ To date, two earlier studies using TR in children with CP have shown promising results in terms of feasibility and effectiveness compared with standard face-to-face (FtF) interventions, however, these studies have not reported objective outcomes.^14,15^

Low-cost virtual reality exergames are well-accepted by children and adolescents due to their ludic, motivating and engaging features, which facilitate a nearly seamless incorporation into TR.^16^ However, research on TR using low-cost virtual reality is yet in its infancy as high-level evidence comparing this modality with standard FtF interventions is still lacking. It is noteworthy that low-cost virtual reality TR also brings the possibility to standardize rehabilitation interventions, which is a pending task in neurorehabilitation.^18^

This clinical trial protocol seeks to establish the feasibility and effectiveness of a rehabilitation programme using low-cost virtual reality aimed at improving postural control in children with CP. It also seeks to compare low-cost virtual reality under two delivery modalities, telerehabilitation (TR) and face-to-face (FtF, control group). We hypothesize that a rehabilitation programme using low-cost virtual reality will improve postural control in children with CP and that this programme delivered using TR will be as effective as a FtF modality. The TR programme is designed to expand the coverage of physiotherapy services for children with CP in low-resource settings and in remote areas, but also to provide access to physiotherapy during restricted mobility as in the case of recent COVID-19-related lockdowns.

## Materials and methods

### Funding

This study was supported by the Innovation Fund for Competitiveness, Maule Regional Government – Chile No. 30.481.923-0.

### Design

This is a double blinded, randomized, controlled trial designed following the Standard Protocol Items: Recommendations for Interventional Trials (SPIRIT), (figure 1).^19^

**Figure 1:**
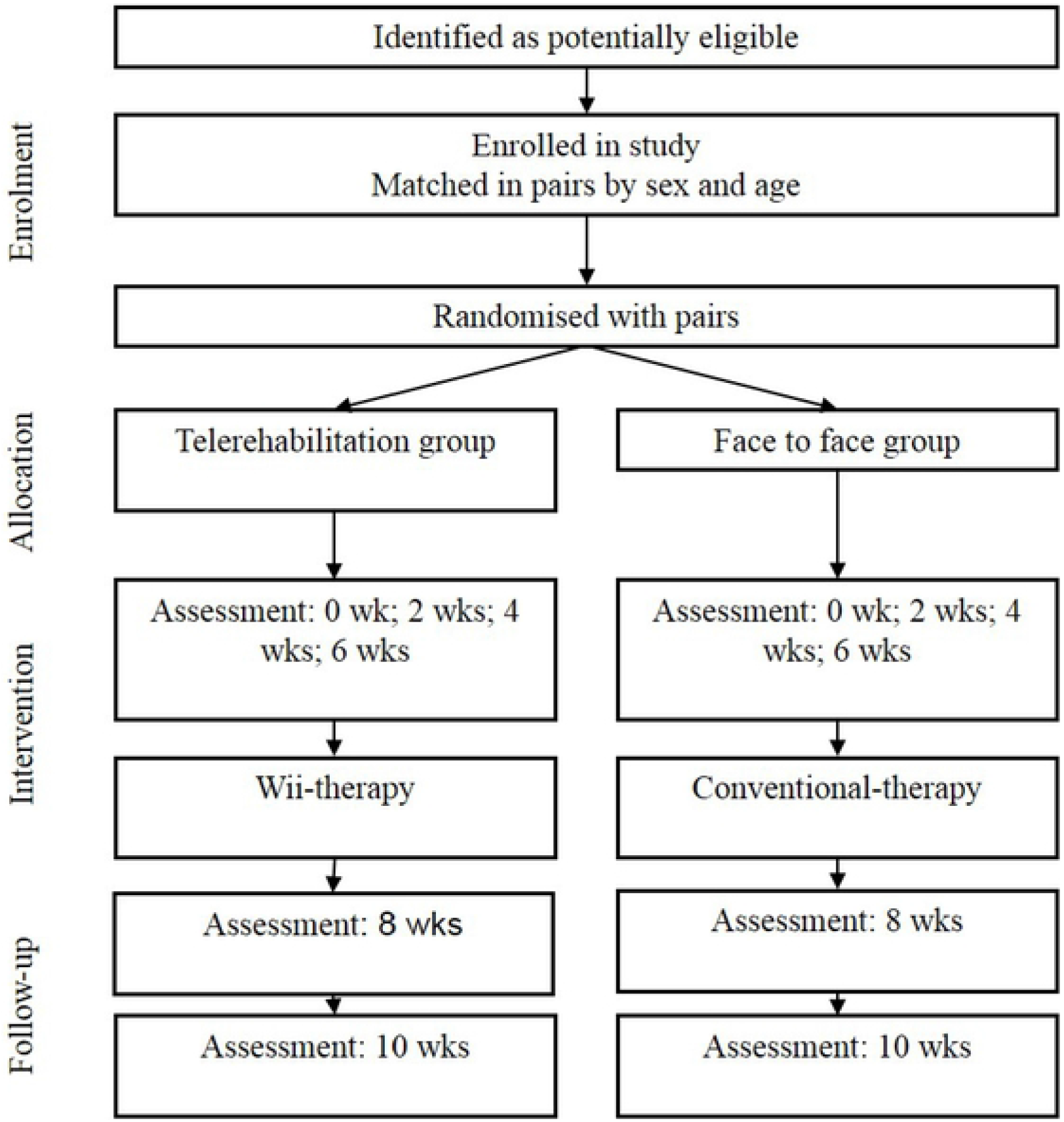
Flow chart according to the Standard Protocol Items: Recommendations for Interventional Trials (SPIRIT). Participants, assessments and interventions. Through the two groups of the trial.

The trial adheres to the Declaration of Helsinki and the Chilean laws of rights and duties of the patient and research in humans. Ethical approval was obtained from the Ethics Committee of the University of Talca (Ref. No. 24-2018). This study was registered with the Australian and New Zealand Clinical Trials Registry (ACTRN12621000117819). Written informed consent will be obtained from parents and participants over 7 years old; verbal assent will be obtained from younger children.

### Study setting

This study compares the effects of low-cost virtual reality in two distinct settings. The first setting is clinical (Telerehabilitation Technology Centre and Neurosciences in Human Movement, Universidad de Talca, Talca, Chile) and in which low-cost virtual reality is delivered in a FtF modality (control group) with therapist interacting directly with the patient. The second setting is a remote facility with no therapist on-site for the delivery of low-cost virtual reality through TR, and could be an at-home, school, or rehabilitation centre. The choice of setting will depend on each participant’s adequate access to space and equipment. All assessments will be conducted at the facility in which therapy was conducted.

### Sample size

The sample size was calculated to detect clinically relevant changes in postural control after low-cost virtual reality in both groups by the end of week 6. Using data from earlier studies,^11,28^ we considered a standard deviation of 4.974 cm^2^ for the CoP sway area (CoP_Sway_), an alpha level of 0.05, 80% statistical power, and an allowance for 5% attrition; we required a minimum of 20 participants in each group (total sample of 40).

### Eligibility criteria

The inclusion criteria are as follows: (1) levels I or II on the Gross Motor Function Classification System (GMFCS)^20^ and on the Expanded and Revised Gross Motor Function Classification System (GMFCS-ER)^21^; (2) aged 7 to 14 years (both sexes) with CP type spastic hemiplegia; and (3) having mild or no cognitive impairment. Exclusion criteria includes patients with other than CP neurological disorders, epilepsy, and uncorrected vision or hearing. Participants with access to a Nintendo Wii at home before the intervention commences will be also excluded.

### Interventions

This study will compare the effects of low-cost virtual reality delivered through FtT and TR modalities. The low-cost virtual reality will consist of in using a Nintendo Wii balance board to play 4 exergames. In the FtF group therapy will be provided by a physiotherapist with experience in the use of exergames for rehabilitation. In the TR modality, the intervention (low-cost virtual reality) will be delivered by the patient’s parent or caregivers. In total, both groups (FtF and TR) will receive 18 sessions of low-cost virtual reality divided in 3 sessions per week (Monday, Wednesday, and Friday) over a period of 6 weeks. Each session will last for 25 minutes in which each participant will perform three series of exergames resting 2 minutes in between. The first two series will include Snowboard, Penguin Slide, and Super Hula Hoop. The first series is played with arms on the side and the second one with hands on the waist to avoid the potential effect of training under one or the other strategy. The third series is deep breathing in the Yoga game. During the first 2 weeks of training (first 6 sessions), patients will receive manual guidance and verbal instructions after which (7^th^ session onwards) only verbal instructions will be provided by the parents, caregiver, or physiotherapist.

In the Snowboard exergame patients displace their centre of pressure (CoP) in the anterior-posterior direction to swerve around flags whereas in the Pinguin Slide movement occur on the medial-lateral direction as patients need to score by catching fishes while maintaining a pinguin inside a continuously tilting platform. Finally, the Hula Hoop consists in maintaining hoops on the waist while moving the CoP in a clockwise and anticlockwise circular pattern. In the Yoga game the aim is to stand as quite as possible first with eyes open and then with the eyes closed

This low-cost virtual reality programme is based on previous studies that have demonstrated similar interventions can effectively train overall postural control.^11^ For all participants in both groups (FtF and TR), adherence will be encouraged through telephone reminders. No modifications of the intervention protocols throughout the study are considered at this stage.

### Outcomes

Differences between the effects of FtF and TR using low-cost virtual reality on postural control will be assessed using posturographic measures. In brief, posturography assesses postural control through measures of the centre-of-pressure sway, of which the most commonly used are sway area and trajectories and velocity; total and in the medial-lateral and anterior-posterior directions.^22^ These CoP variables have been shown to be sensitive in determining changes in postural control due to training in young adults,^23^ patients with post-stroke hemiparesis,^24^ patients with Parkinson’s disease,^25,^ and children with CP.^26-30^

Primary outcome in this study will be the CoP sway area (CoP_area_), which has been shown to be a reliable and valid measure of postural control in different clinical and nonclinical populations.^22-30^ The CoP_area_ is an overall measure of the ability of the balance control system to maintain a stable upright posture, to note, higher CoP_area_ values indicate poorer balance control.

Secondaries outcome measures from the posturographic assessments to be used in this study will be: standard deviation and velocity of the CoP in the medial-lateral and anterior-posterior directions. The CoP standard deviation is a measure of the variability of CoP displacements whereas the CoP velocity is a measure of the ability to react to changes in posture with larger values indicating poorer balance control. The latter is due to late postural adjustments lead to large CoP excursion that demand rapid corrections to maintain stability.^22^

Tertiary outcomes of this study will be: the Modified Modified Ashworth Scale (MMAS) for lower limbs (ankle and knee flexor-extensor musculature), Modified Ashworth Scale (MAS) for upper limbs (elbow and wrist flexor-extensor musculature), timed up-and-go (TUG) tests, the timed one-leg standing (TOLS) and 6-minute walk test (6MWT). These are widely used clinical measures of spasticity (MMAS and MAS) and overall physical performance, which include some aspects of postural control (TOLS) and dynamic balance (TUG and 6MWT).

### Intervention timeline

Participants will be assessed at baseline, by the end of weeks 2, 4, and 6, and at weeks 8 and 10 (post-intervention follow-ups) using posturography and clinical measures.

### Recruitment

Children and adolescents with congenital SHE-CP will be recruited from the Rehabilitation Institute, Talca, Chile. This is an outpatient clinic that provides rehabilitation services to patients with neurological disorders including CP children. It is noteworthy, that patients from this centre will have little or no experience using a Nintendo Wii balance board. An experienced staff of health professionals, comprised of a physician and two physical therapists, will recruit participants according to the inclusion/exclusion criteria. Screening for potential participants will continue until the calculated sample size is achieved.

### Allocation

Participants will be randomly assigned to either control (FtF) or experimental group (TR) with a 1:1 allocation as per a computer-generated randomization schedule stratified by site and the baseline score of the Action Arm Research Test (ARAT; <= 21 versus >21) using permuted blocks of random sizes. The block sizes will not be disclosed, to ensure concealment.

### Management

Participants will be informed of the group they will be randomly assigned to at the beginning of the study. Participants in each group will be systematically asked about the content of the interventions received to maintain engagement and compliance. Due to the nature of the intervention neither participants nor staff can be blinded to allocation, but are strongly inculcated not to disclose the allocation status of the participant at the follow up assessments. An employee outside the research team will feed data into the computer in separate datasheets so that the researchers can analyse data without having access to information about the allocation.

All raw data will be electronically saved in coded text files whereas calculated and clinical outcomes measures will be saved in a coded Excel spreadsheet. Data backups will be made every week.

### Data collection

All measurements will be performed at the same time of the day, under the same conditions and by the same assessor at each time point. Posturographic measures (CoP displacements) for both groups will be collected using an AMTI OR67 force platform (Watertown, MA, USA). The AMTI NetForce software (Watertown, MA, USA) will be used for the acquisition of moments and forces at 200Hz Data will be recorded, coded, and stored on a personal computer. CoP variables will be calculated in Matlab R2012 (Mathworks Inc., Natick, MA, USA).

All primary and secondary posturographic measures will be assessed under 2 static and 4 dynamic conditions for 30 seconds each: (i) standing still with eyes open, (ii) standing still with eyes closed, voluntary sway in the mediolateral direction following a metronome set at (iii) 30Hz and (iv) 60Hz, and multidirectional sway while playing 2 different videogames that challenge (v) anterior-posterior postural control (snowboard) and (vi) medial-lateral postural control (pinguin). It is expected that all assessments, including clinical ones, will be completed within 30 minutes.

### Data analysis

Statistical analyses will be performed using SPSS 20.00 for Windows. Descriptive statistics will be used to calculate the demographic and clinical characteristics of participants in both, FtF and TR groups (unpaired t-tests and χ² tests).

For all outcome measures, the Shapiro–Wilk and Levene tests will be used to measure the normality and homogeneity of the variance, respectively. The Mann–Whitney U test will be used to determine differences between the FtF and TR groups. Friedman’s one-way analysis of variance (ANOVA) with post hoc pairwise comparisons (Wilcoxon signed-rank test) will be used to determine the effect over time for each group (FtF and TR). For all analyses, a *p*-value ≤0.05 will be considered statistically significant.

### Data monitoring

A permanent monitoring will be made by the clinical team, oversighted by the research team and the Ethics Committee of the University of Talca. All the possible adverse events will be informed immediately to the Ethics Committee of the University of Talca to determine trial’s modifications or stop.

## Discussion

The aims of this clinical trial protocol are to establish the feasibility and effectiveness of a low-cost virtual reality to improve postural control in children with CP and to compare two low-cost virtual reality delivery modalities, telerehabilitation (TR) and face-to-face (FtF). Demonstrating that a TR is as efficient as a FtF intervention is crucial to adopt TR as an alternative to expand the coverage of rehabilitation services for children with CP in low-resource settings and in remote areas. Further, by showing the efficacy of TR as a method to deliver low-cost virtual reality, physiotherapy can be potentially delivered when services are inaccessible or closed as in the case of recent covid-19-related lockdowns. In this protocol we hypothesize that low-cost virtual reality will improve postural control and that both delivery methods will be equally effective. The latter is reasonable considering that our previous research has shown that the FtF modality is effective in improving static and dynamic postural control and reducing spasticity in children and adolescents with spastic cerebral palsy.^29-32^.

High-intensity physiotherapy is necessary for optimizing motor learning and improve physical skills in patients with CP.^33^ Regrettably, children and adolescents with CP typically receive limited physiotherapy sessions, which implies a lost opportunity to improve the functionality and quality of life of these patients during a critical period of their lives. It is important to note that the neural maturity of sensory systems responsible for the control of postural balance is sequential and dependent on age, reaching full maturity at 15-16 years.^34, 35^ Therefore, providing postural control rehabilitation during this age period is essential to generate changes in the lives of these patients that will accompany them into adulthood.

There are several factors that affect standing balance: not only the type of CP but also other medical and environmental conditions. This diversity habitually demands for tailored therapy that is not easily accessible, especially in low-resource and/or rural settings. Then, this heterogeneity is consistent with the plethora of interventions available that are hard to standardize precisely because of the diversity of patients, their needs, environments, and resources, among other aspects^36-39^.

Virtual reality and low-cost virtual reality could alleviate this tension by creating a plethora of programs that can be replicated in different settings in a tailored way.

In this regard, virtual environments created by software are characterized as artificial functional spaces of multi-sensory interaction with a motivational purpose and are easy to manipulate.^40^ These virtual environments can be tailored for each patient, but all these modifications are transferable for other patients and settings; this eventuality provides the possibility to standardize most of the rehabilitation therapy. Furthermore, the standardization of virtual reality-based therapies seems to be related to the adoption of commercial video games,^41,42^ which are widely accessible worldwide. However, this does not guarantee that these kinds of therapies are really standardized. The number of sessions, duration, sequence, games, intervals, and other aspects need to be defined to obtain a transferable standard. A recent systematic review on the effectiveness of virtual reality in children and young adults with CP found limited confidence in effect estimation for utilising virtual reality in this population; thus, further research is necessary.^18^

A low-cost virtual reality device provides greater accessibility and transferability to patients with CP at neurological care centers. The Nintendo Wii system offers a simple and affordable mode of virtual reality therapy.^43,44^ The Nintendo Wii and its peripheral balance board have been proposed as a training tool to improve standing balance in the elderly, adults with total knee replacements, post-stroke patients, and patients with Parkinson’s disease.^9,45-48^

Another important consideration to generate relevant evidence is comprehensive measurements. Thus, evaluations to build postural control construct and to determine the effects of physical interventions in these populations must include clinical and laboratory measurements. This clinical trial includes a set of clinical and laboratory evaluations that enable knowing the effects of these therapies in the different aspects of postural control, for example, the levels of the sensory systems—visual, vestibular, and mechanoreceptive systems—in static and dynamic motor skills, within them include walking.

Home-based therapies for patients with CP seem to be feasible; however, the available studies have used diverse methods and strategies, and, therefore, it is hard to determine the impact of these kinds of therapies.^49^ An efficient way to deliver these home-based therapies is using TR, which appears to be an option to improve physical function in patients with disabilities. However, most research has included low-quality evidence studies that make it impossible to determine the effectiveness of TR.^50^ In the case of patients with CP, two studies have shown promising results,^13,14^ but the evidence is far from conclusive.

We have selected posturographic measures that have shown to be sensitive to the effects of physiotherapy interventions.^51-52^ Velocity of CoP describes the neuromuscular response to shifts in the body’s centre of mass and serves as an indicator of stability. The mean velocity of CoP has high reliability (ICC > 0.8) as a measurement parameter during standing balance and is sensitive to changes of age-related postural impairment.^52^ However, the area of the CoP presents behaviour that is more difficult to modify after physical interventions in physiotherapy. For the area of the CoP (CoP_Sway_), greater values indicate poorer balance control as CoP gets closer to the limit of stability, which demands rapid stabilization responses expressed in greater CoP velocities (V_ML_ and V_AP_). Generating a modification in postural control, specifically reducing the CoP area, involves working exhaustively at a dose capable of achieving it. The dose in this study has been reviewed and verified both with the scientific literature and tested in previous research.^28^ There are correlation studies between posturographic platform variables and clinical variables. In line with this, the area of the CoP and other variables are poorly correlated with the clinical variables.^53,54^ These findings suggest that the dose of physical therapies should be constantly reviewed, and second, that clinical variables are more difficult to be transformed after a physical intervention. This is related to the interpretation of the patients themselves in the face of physical changes that their body and motor control has. There is evidence that the perceptions of better postural control, balance, static, and dynamic stability by patients are conditioned by the positive changes in the clinical variables investigated in these evaluations. Therefore, it is essential to know the effects of this type of virtual physical therapy in clinical trials and to consider pertinent clinical and gold standard laboratory evaluations, such as the force platform, both of which will build the postural control construct and interpret the results towards the patients.

Despite being discontinued in 2014, the Nintendo Wii continues to be a relevant tool for neurorehabilitation research. Recent research using this console to treat patients with CP has shown positive effects compared to other kinds of rehabilitation services.^55-57^

This will be the first study assessing the effects of a six-week exercise programme remote (TR) using a Nintendo Wii balance board to improve standing balance in children and adolescents with CP type spastic hemiplegia compared with the effects of an FtF modality. This trial will provide evidence on the effectiveness of the Nintendo Wii for improving standing balance in patients with CP, a matter in which the current evidence is lacking. This low-cost virtual reality TR programme is designed to expand the coverage of physical therapy services for patients with CP in low-resource settings using affordable products, such as the Nintendo Wii balance board. The scientific contribution towards improving static or dynamic standing balance will support the idea that affordable technological tools can positively contribute to the usual activities, such as walking, running, and climbing stairs, that children with CP perform in their environment, namely playgrounds, schools, and in the cities where they live. During the pandemic, the development of new technological strategies in physical therapies has contributed to the advancement in rehabilitation, this should allow in the future to install new proposals in physical rehabilitation for the children with CP type spastic hemiplegia.

## Data Availability

Data cannot be shared publicly because of Law 20,584 of "Rights and duties of the patient" in Chile. Data are available from the Ethics Committee (contact via, Mr Javier barra) for researchers who meet the criteria for access to confidential data.

